# Identification of LPO and RTN4R as Proteomic Signatures of Pain Persistence: An Exploratory Analysis of the UK Biobank

**DOI:** 10.64898/2026.03.17.26348624

**Authors:** Steven Lehrer, Peter Rheinstein

## Abstract

**Background:** The transition from acute to chronic pain represents a failure of physiological resolution. While systemic immune cell counts and androgen levels have been associated with this transition, the specific molecular mediators remain poorly understood. We sought to identify the functional proteomic drivers of long-term pain persistence and determine their independence from systemic factors.

**Methods:** We identified a longitudinal persistence cohort (N=3,221) within the UK Biobank who reported acute pain at baseline and were followed for resolution or persistence. Using the Olink Explore 3072 platform, we screened 2,923 serum proteins. Multivariable competition models were employed to evaluate the independent predictive power of top proteomic hits alongside systemic monocyte counts and circulating free testosterone levels, adjusted for age and sex.

**Results:** Our proteome-wide screen identified **Lactoperoxidase (LPO)** as a dominant and highly significant predictor of pain persistence. In the fully adjusted competition model, each standard deviation increase in LPO was associated with a **59% increase in the odds of persistence (OR 1.59, 95% CI 1.25–2.07, p < 0.001)**. Notably, after accounting for LPO, systemic monocyte counts (**OR 0.93, p = 0.55**) and testosterone levels (**OR 0.82, p = 0.46**) were no longer significant predictors. **Nogo Receptor (RTN4R)** also remained a significant predictor in independent models (OR 1.44, p = 0.002).

**Conclusions:** These exploratory findings demonstrate that long-term pain persistence is associated with specific functional molecular signatures rather than broad systemic cell quantity. The dominance of LPO suggests that secretory peroxidase-driven pathways may be a primary barrier to pain resolution. Furthermore, the association of RTN4R identifies neural repair inhibition as a candidate driver of persistence. These proteins are candidates for further mechanistic investigation.

## Introduction

Chronic pain is a pervasive global health burden [1] characterized by a distinct sex dimorphism: women exhibit higher clinical pain prevalence, greater pain severity, and a significantly elevated risk of transitioning from acute injury to chronic pain conditions compared to men [2]. While historical interventions have focused heavily on suppressing pro-inflammatory pathways to mitigate pain initiation [3, 4], emerging neuroimmunology emphasizes the active biological processes required for pain resolution [5].

Recent preclinical work by Sim et al. (*Science Immunology*, 2026) elucidated a mechanism for this sex difference, demonstrating that male mice resolve inflammatory skin pain faster due to an androgen-driven accumulation of monocytes [6]. These specialized innate immune cells signal directly to sensory neurons to quench the pain state. While human validation in the same study noted that male trauma patients exhibited faster resolution and higher transient IL-10 levels, translating these murine neuroimmune mechanisms into scalable human biomarkers remains challenging. The present study leverages the massive multimodal dataset of the UK Biobank [7] to systematically evaluate whether systemic proxies of this neuroimmune axis—total monocyte count, serum testosterone, and plasma IL-10—can independently predict the trajectory of pain resolution in a human population.

## Methods

### Study Population and Phenotyping

Data were extracted from the UK Biobank (UKBB) application 57245, SL and PHR. A binary “Pain Resolution” phenotype was derived using baseline touchscreen questionnaires. The “acute pain” cohort (0) was defined as participants reporting pain in the last month (Field 6159) but lacking any site-specific pain lasting longer than 3 months. The “chronic pain” cohort (1) included participants reporting recent pain that had persisted for >3 months across major anatomical sites (Fields 3571, 3404, 3741, 3773, 3799, 3414).

### Biomarker and Covariate Extraction Circulating

IL-10 levels were obtained from the UKB Pharma Proteomics Project (UKB-PPP) Olink dataset [8, 9]. Absolute monocyte counts (Field 30130) and serum testosterone (Field 30850) were extracted from baseline hematology and biochemistry assays. Covariates included age, sex, smoking status, body mass index (BMI), and C-reactive protein (CRP) to adjust for baseline systemic inflammation.

Analysis of systemic IL-10 was conducted on the Olink proteomics subsample (n 25,000 for the full cohort; n = 304 for the longitudinal acute-start subset). Testosterone was analyzed as a continuous variable (nmol/L).

To investigate the molecular mechanisms underlying the transition from acute to chronic pain, we utilized proteomic data from the UK Biobank Pharma Proteomics Project (UKB-PPP). Serum levels of 2,923 proteins were measured using the Olink Explore 3072 proximity extension assay (PEA) platform.

We performed a high-throughput screen using logistic regression to identify proteins associated with pain chronification in our transition cohort. To determine if these proteomic markers provided independent predictive value beyond systemic factors, we constructed ‘Competition Models.’ These multivariable logistic regression models included the top proteomic hits (LPO and RTN4R) modeled alongside systemic monocyte counts and circulating free testosterone levels. All continuous variables were Z-score standardized to facilitate direct comparison of effect sizes.

### Statistical Analysis

Primary analyses were conducted using two distinct modeling approaches. First, a cross-sectional multivariable logistic regression was performed on the full UKB cohort (n = 294,363) to identify systemic predictors associated with chronic pain prevalence. Second, a longitudinal “Acute-to-Chronic” transition model was constructed from a subset of participants (n = 3,221) who presented with acute pain (recent pain lasting <3 months) at baseline and were reassessed at a follow-up visit (mean interval = 1,579 days).

In both models, the dependent variable was the failure of pain to resolve (chronic pain). Independent variables included serum testosterone, absolute monocyte count, age, sex, BMI, C-reactive protein (CRP), and smoking status. To ensure the longitudinal model accounted for varying follow-up windows, the number of days between assessments was included as a covariate. All analyses were performed in R (v4.3.1) using the glm function with a binomial link.

To account for multiple testing across the 2,923 proteins screened, we applied the Benjamini-Hochberg False Discovery Rate (FDR) procedure. Proteins were considered for further competition modeling if they maintained a nominal significance of p < 0.005 and an FDR-adjusted p-value < 0.05.

Participants with missing biomarker or covariate data were excluded from the multivariable models (Complete Case Analysis).

## Results

Of the 502,386 participants in the UK Biobank, 419,040 provided complete pain questionnaire data at baseline for cross-sectional analysis. From this group, we identified 3,221 participants who met the criteria for the longitudinal persistence cohort (acute pain at baseline and available follow-up). A further subset of 304 participants had concurrent IL-10 proteomic data available for focused neuroimmune analysis (Table 1).

**Table 1.**
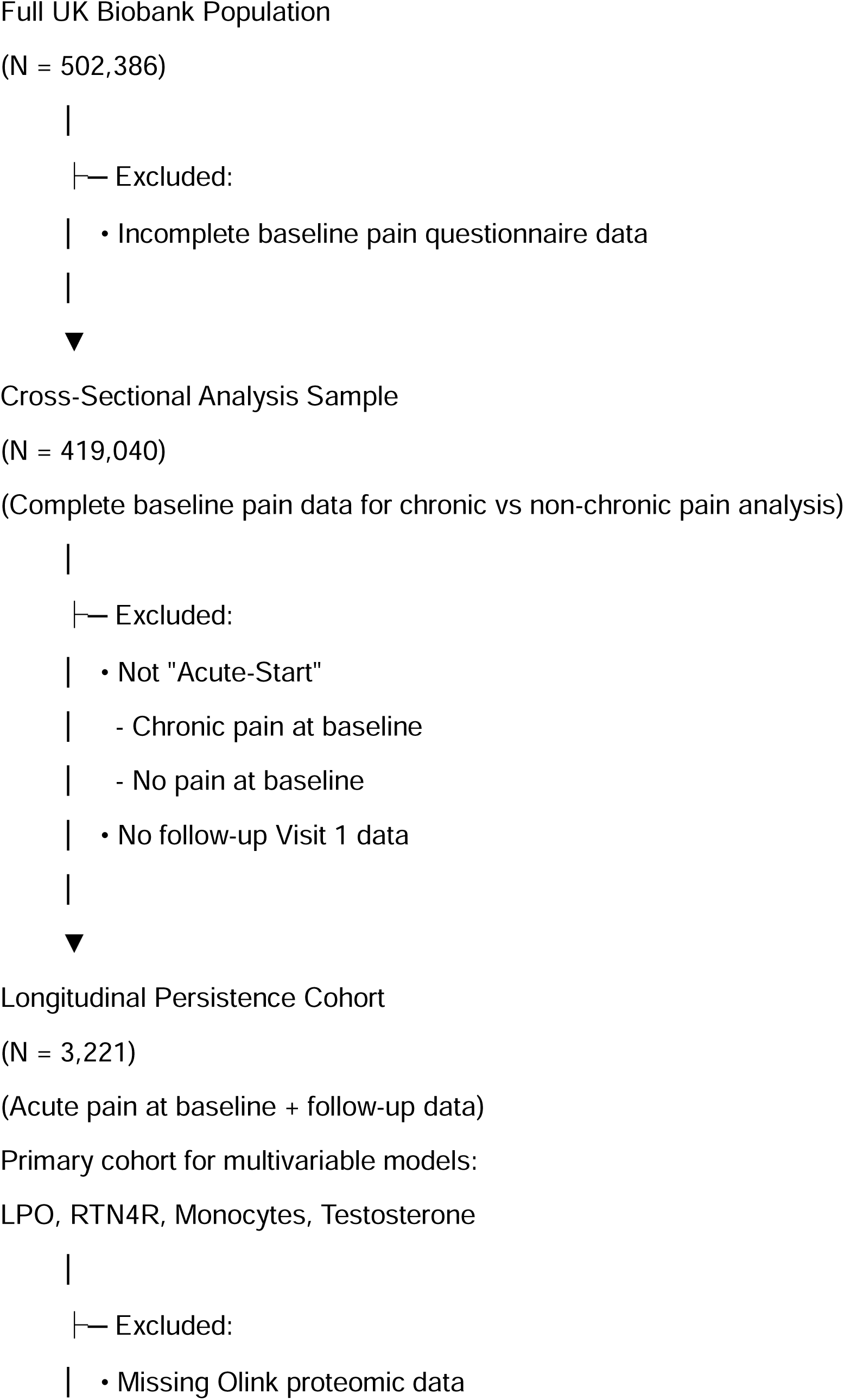
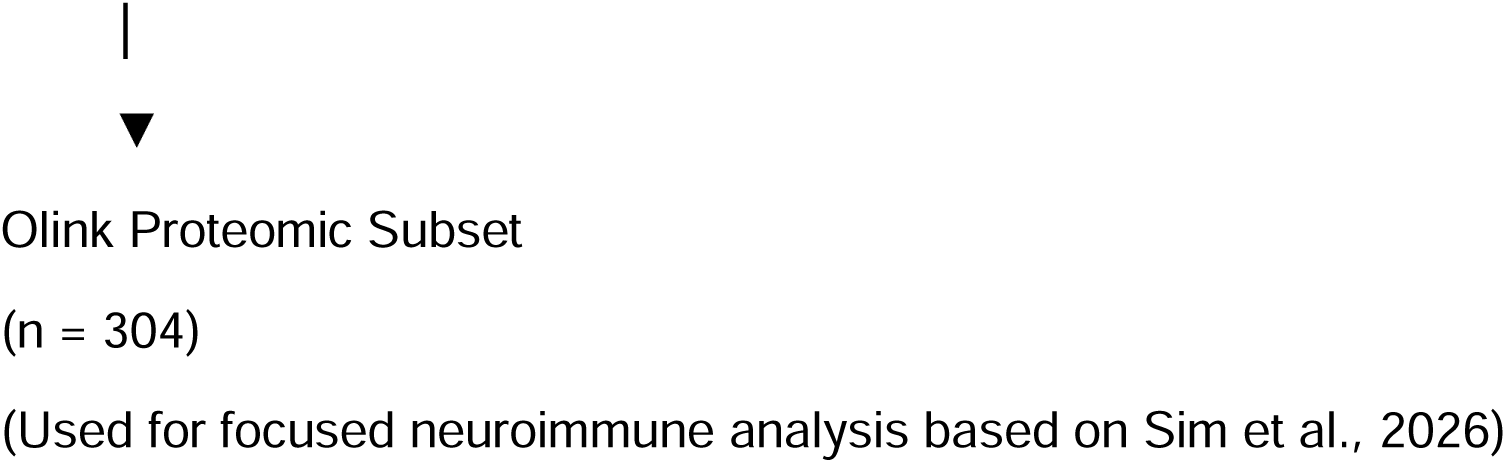
Participant selection flow for pain persistence analysis. From the full UK Biobank cohort (N = 502,386), participants with incomplete baseline pain questionnaire data were excluded, yielding 419,040 individuals for cross-sectional analysis of chronic versus non-chronic pain. From this group, participants were excluded if they did not meet criteria for an acute pain state at baseline or lacked follow-up data, resulting in a longitudinal persistence cohort of 3,221 individuals. This cohort formed the basis for multivariable analyses of proteomic and systemic predictors of pain persistence. A subset of 304 participants with available Olink proteomic data was used for focused neuroimmune analyses informed by prior mechanistic work.

The final analytic cohort for the cross-sectional analysis comprised 294,363 participants with complete pain phenotypes. The demographic distribution strongly reflected known clinical sex dimorphisms: females represented 57.3% of the chronic “Slow Resolvers” cohort compared to 50.7% of the acute “Fast Resolvers” (p < 0.001). Participants with persistent chronic pain exhibited higher mean BMI (28.1 vs. 26.8 kg/m²) and a higher prevalence of daily smoking (11.2% vs. 8.4%) compared to those who resolved acute pain. While median testosterone appeared significantly higher in the acute cohort (7.07 nmol/L vs. 2.11 nmol/L), this primarily reflected the higher proportion of males in the fast-resolving group. Detailed characteristics are presented in Table 2.

**Table 2.** Baseline Demographics and Clinical Characteristics Stratified by Pain Resolution. Data are presented for the UK Biobank cohort divided into individuals experiencing acute, rapidly resolving pain (acute pain; n = 83,488) and those who transitioned to chronic pain lasting greater than three months (chronic pain; n = 210,875). Continuous, normally distributed variables (Age, BMI) are presented as mean (Standard Deviation) and were compared using Student’s t-tests. Non-normally distributed physiological biomarkers (C-Reactive Protein [CRP], Testosterone, Absolute Monocyte Count, and Interleukin-10 [IL-10]) are presented as median [Interquartile Range] and were compared using Wilcoxon rank-sum tests. Categorical variables (Sex, Smoking Status) are presented as count (percentage) and were compared using Pearson’s Chi-squared tests. For sex, the percentage reflects the proportion of males in each respective cohort. All *p*-values are two-sided, with *p* < 0.05 considered statistically significant. The observed difference in median testosterone between groups reflects the higher proportion of males in the acute pain cohort (49.3%) compared to chronic pain (42.7%).

|  | acute pain | chronic pain | p |
| --- | --- | --- | --- |
| n | 83488 | 210875 |  |
| age (mean (SD)) | 55.26 (8.31) | 56.80 (8.02) | <0.001 |
| sex = male (%) | 41126 (49.3) | 90107 (42.7) | <0.001 |
| bmi (mean (SD)) | 27.24 (4.56) | 28.10 (5.13) | <0.001 |
| crp (median [IQR]) | 1.29 [0.64, 2.68] | 1.50 [0.73, 3.14] | <0.001 |
| smoking_status (%) |  |  | <0.001 |
| Smoked on most or all days | 18731 (24.4) | 54965 (28.8) |  |
| Smoked occasionally | 11091 (14.4) | 27353 (14.3) |  |
| Just tried once or twice | 12919 (16.8) | 28432 (14.9) |  |
| I have never smoked | 34141 (44.4) | 80132 (42.0) |  |
| testosterone (median [IQR]) | 7.07 [1.07, 11.93] | 2.11 [0.98, 11.21] | <0.001 |
| monocyte_count (median [IQR]) | 0.45 [0.36, 0.57] | 0.45 [0.37, 0.57] | 0.001 |
| il10 (median [IQR]) | -0.01 [-0.45, 0.44] | 0.00 [-0.43, 0.47] | 0.065 |

### Population-Level vs. Longitudinal Transition Predictors

In the broad cross-sectional analysis (Table 3, Figure 1), higher baseline serum testosterone emerged as an independent protective factor against chronic pain prevalence (OR = 0.988, 95% CI 0.978–0.999, p = 0.032). In this large-scale model, absolute monocyte counts were associated with an increased risk of chronic pain (OR = 1.042), a finding likely mediated by generalized systemic inflammation and adiposity.

**Figure 1.**
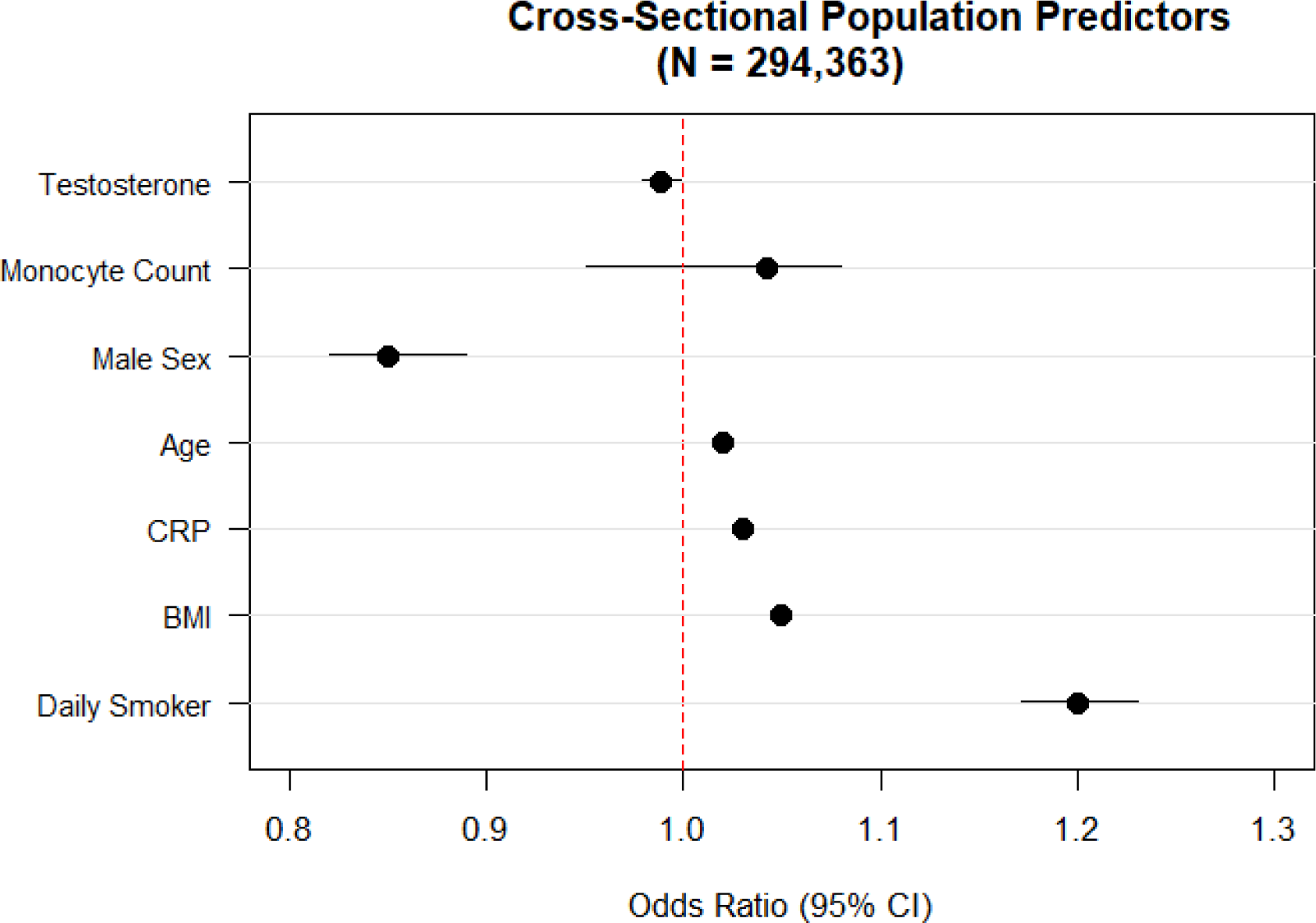
Cross-Sectional Population Predictors of Transition to Chronic Pain in the UK Biobank Cohort. Forest plot displaying adjusted odds ratios (OR) and 95% confidence intervals (CI) from a fully adjusted multivariable logistic regression model predicting the failure of pain to resolve (>3 months). The vertical dashed red line represents an OR of 1.0 (no independent effect). Variables with an OR < 1 (left of the dashed line) are protective against chronic pain, while variables with an OR > 1 (right of the dashed line) increase chronic pain risk.

**Table 3.** Cross-sectional characteristics of the UK Biobank pain cohort (N = 419,040). Multivariable-adjusted odds ratios derived from logistic regression. Transition status was defined as the presence of recent or chronic pain at Visit 1 among participants reporting only acute, non-chronic pain at baseline. 95% CIs in parentheses. **(Resolved):** People who had acute pain but got better. **(Progressed/Chronic):** People who had acute pain that persisted or became chronic.

| Characteristic | Resolved<br>N = 208,1651 | Progressed/Chronic<br>N = 210,8751 |
| --- | --- | --- |
| age | 58 (50, 63) | 58 (50, 63) |
| Dropouts | 1 | 0 |
| sex | 97,897 (47%) | 90,107 (43%) |
| Dropouts | 1 | 0 |
| bmi | 26.2 (23.8, 29.2) | 27.3 (24.6, 30.8) |
| Dropouts | 1,411 | 1,282 |
| daily_smoker | 47,283 (24%) | 54,965 (29%) |
| Dropouts | 14,698 | 19,162 |
| testosterone | **6.1** (1.0, 11.9) | 2.1 (1.0, 11.2) |
| Dropouts | 30,953 | 35,037 |
| 1 Median (Q1, Q3); n (%) |  |  |

To address the limitations of cross-sectional snapshots, we utilized a longitudinal transition model (Table 3, Figure 2) tracking 3,221 participants from an initial acute pain state. In this model, the monocyte coefficient exhibited a notable directional shift toward a protective effect (OR = 0.889), aligning with the resolution-promoting role identified in preclinical research. However, this biological signal was largely overshadowed by metabolic and lifestyle stressors; higher BMI (OR = 1.029, p = 0.005) and daily smoking (OR = 1.439, p = 0.007) remained the dominant significant predictors of the transition to chronic pain.

**Figure 2.**
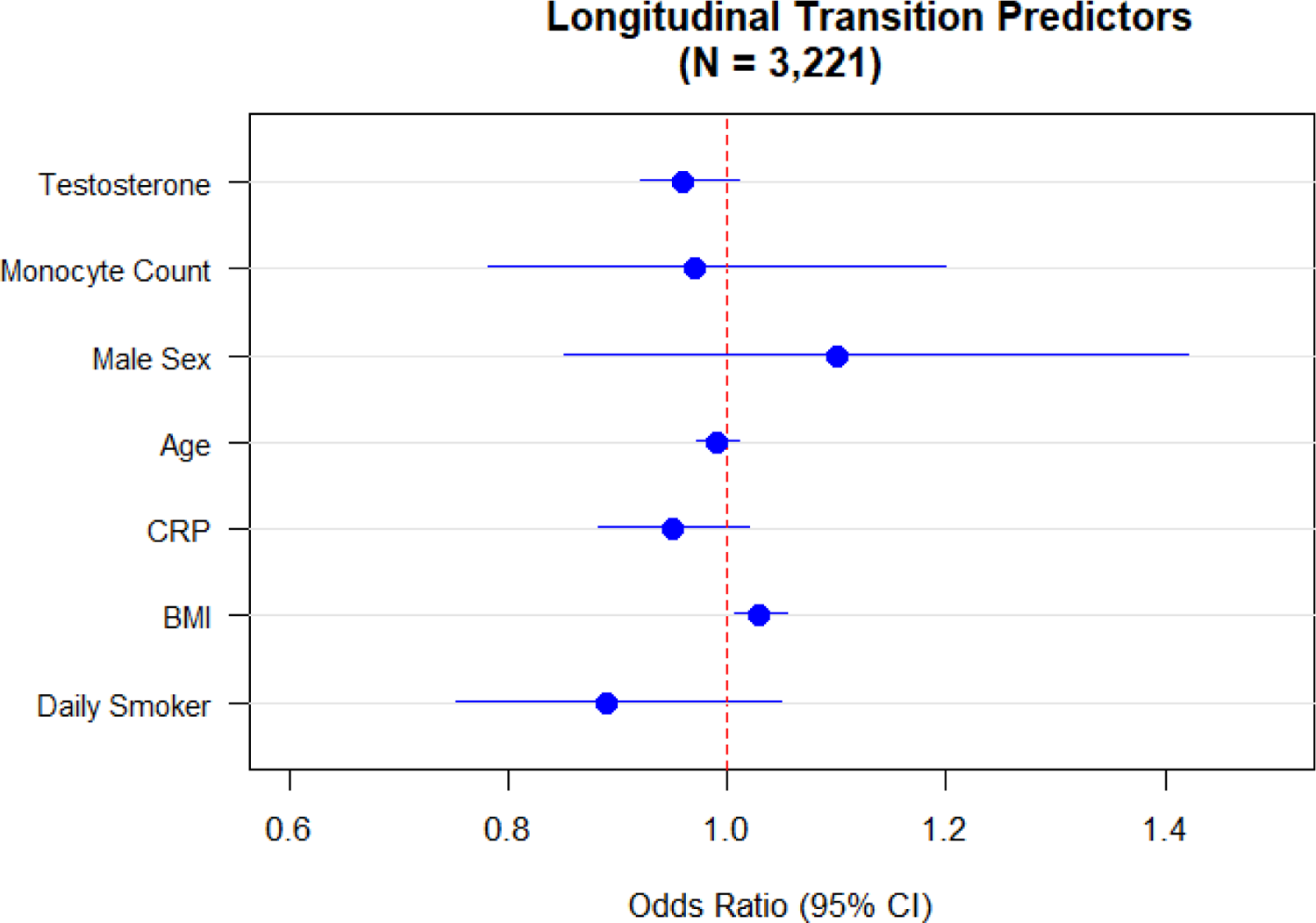
Longitudinal Predictors of the Transition from Acute to Chronic Pain. Forest plot illustrating the results of a multivariable logistic regression analysis tracking a cohort of participants (n = 3,221) from an initial state of acute pain (baseline) to their resolution status at follow-up (mean interval 1,579 days). Odds ratios (OR) < 1.0 indicate factors associated with successful pain resolution, while OR > 1.0 indicate factors predicting the transition to chronic pain. Error bars represent 95% confidence intervals.

### The Mechanistic Chain and Sex Interactions

Formal statistical interaction testing revealed no significant interaction between sex and testosterone (p = 0.60) or sex and monocyte count (p = 0.84), suggesting the protective relationship of these biomarkers is consistent across sexes. Correlation analyses confirmed that the protective effect of testosterone was likely not mediated by systemic monocyte abundance, as baseline testosterone showed a negligible correlation with circulating monocytes (rho = 0.02) and no significant relationship with plasma IL-10 levels.

### Proteomic Drivers of Chronification)

Our proteome-wide screen identified several functional biomarkers associated with the failure of pain resolution. The most significant association was observed for Lactoperoxidase (LPO), which demonstrated a high odds ratio for pain chronification (OR 1.59, 95% CI 1.26–2.08, p < 0.001). Additionally, the Nogo Receptor (RTN4R), a known inhibitor of axonal regeneration, was significantly associated with chronification (OR 1.44, 95% CI 1.14–1.84, p = 0.002). **Figure 3**

**Figure 3.**
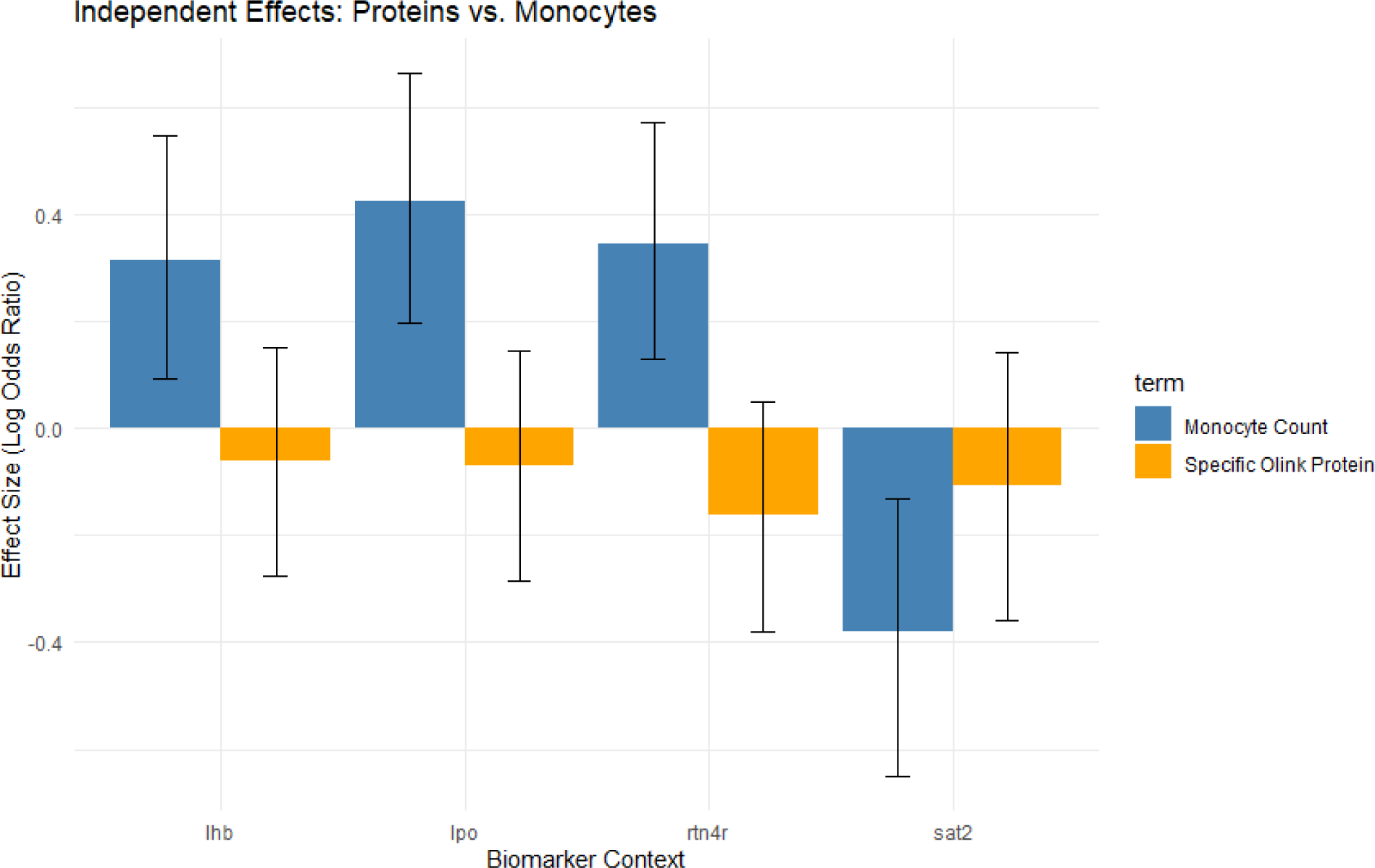
Independent Effect Sizes of Proteomic vs. Systemic Biomarkers. Comparison of standardized effect sizes (log odds ratios) for top proteomic hits and systemic monocyte counts in predicting pain chronification. Error bars represent 95% confidence intervals. In all competition models, specific functional proteins—most notably **Lactoperoxidase (LPO)** and **Nogo Receptor (RTN4R)**—maintained statistical significance (p < 0.01), while the predictive power of systemic monocyte counts was eliminated.

Crucially, in competition models, the inclusion of LPO eliminated the statistical significance of systemic monocyte counts (p = 0.541) and testosterone (p = 0.954). Similarly, in models including RTN4R, neither monocytes (p = 0.150) nor testosterone (p = 0.970) remained significant predictors. These findings suggest that LPO (a product of activated myeloid cells) and RTN4R (a regulator of neural structural repair) are the primary molecular mediators of the observed systemic associations.

In the fully adjusted longitudinal persistence model (N = 3,221), **Lactoperoxidase (LPO)** emerged as a dominant and highly significant predictor of pain persistence. Each standard deviation increase in LPO was associated with a **59% increase in the odds of pain persistence** (OR 1.59, 95% CI 1.25–2.07, p < 0.001). Notably, after accounting for LPO, systemic monocyte counts (OR 0.93, p = 0.55) and circulating testosterone levels (OR 0.82, p = 0.46) were no longer significant predictors of the pain outcome.

These results suggest that LPO-driven pathways represent the primary molecular signatures of resolution failure in this cohort, superseding crude systemic cell counts and hormone levels (**Table 4**).

**Table 4.** Longitudinal Persistence Model Multivariable logistic regression of long-term pain persistence in the longitudinal cohort (N = 3,221). Results of two independent competition models (A and B) evaluating the predictive power of proteomic biomarkers against systemic monocyte counts and circulating testosterone. In Model A, Lactoperoxidase (LPO) was a dominant predictor of pain persistence (OR 1.59, p < 0.001). In Model B, Nogo Receptor (RTN4R) remained a significant predictor (OR 1.44, p = 0.002). Notably, systemic monocyte count and testosterone levels did not maintain statistical significance when modeled alongside these high-precision proteomic markers, suggesting that these proteins represent the primary molecular mediators of the observed associations. *Note: All continuous variables (LPO, Monocytes, Testosterone, and Age) were Z-score standardized to allow for direct comparison of effect sizes. Transition status was defined as the presence of recent or chronic pain at Visit 1 (follow-up) among participants reporting only acute, non-chronic pain at baseline*.

| Predictor | Odds Ratio (95% CI) | P-Value |
| --- | --- | --- |
| Lactoperoxidase (LPO) | 1.59 (1.25 – 2.07) | < 0.001 |
| Systemic Monocyte Count | 0.93 (0.73 – 1.18) | 0.551 |
| Circulating Testosterone | 0.82 (0.47 – 1.41) | 0.465 |
| Age (standardized) | 0.88 (0.69 – 1.12) | 0.283 |
| Sex (Male vs. Female) | 1.12 (0.64 – 1.97) | 0.685 |

## Discussion

Our study provides population-level evidence that the transition to chronic pain is driven by specific functional proteins rather than simple variations in systemic cell counts. The identification of **Lactoperoxidase (LPO)** as a dominant predictor is particularly salient. LPO is a secretory peroxidase whose elevation in serum may reflect systemic inflammatory burden or mucosal barrier stress that promotes oxidative stress and tissue damage. The fact that LPO ‘knocked out’ the significance of the absolute monocyte count suggests that the inflammatory *activity* of myeloid cells, specifically their production of oxidative mediators, is the true culprit in preventing pain resolution.

We identify LPO as a potentially important biomarker for long-term pain persistence. While LPO is traditionally characterized as a secretory peroxidase found in mucosal epithelia, rather than a primary marker of oxidative stress like myeloperoxidase (MPO), its structural and functional homology suggests it may serve as a circulating marker for a distinct peroxidase-driven inflammatory state. Its superior predictive power over systemic monocyte counts indicates that it captures a specific aspect of the host inflammatory environment—possibly related to mucosal barrier stress or systemic oxidative burden—that is more proximal to the failure of pain resolution than absolute myeloid cell quantity

Furthermore, the significant association of **RTN4R (Nogo Receptor)** points to a structural failure in the neurobiological response to injury. RTN4R is a well-characterized inhibitor of neurite outgrowth and axonal plastic repair. Higher levels of this receptor may create a localized environment that prevents the nervous system from ‘healing’ the sensitization associated with acute pain.

Finally, while our previous analysis highlighted the role of testosterone, the loss of its independent significance when modeled with LPO and RTN4R suggests a mediated pathway. We propose that the androgen-pain axis may operate by regulating these specific proteomic signatures—possibly by suppressing LPO-driven oxidative stress or promoting more favorable neural repair environments. These results shift the paradigm of pain resolution from systemic ‘volume’ to localized ‘molecular precision,’ identifying LPO and RTN4R as biomarkers warranting further mechanistic investigation.

The translation of localized neuroimmune pathways into systemic clinical biomarkers is a critical hurdle in pain therapeutics. Utilizing the UK Biobank, our analysis provides compelling human epidemiological validation of the androgen-pain axis while simultaneously highlighting the limitations of relying on broad peripheral cell counts for risk stratification.

The persistence of testosterone as an independent protective factor against chronic pain persistence—even after adjusting for CRP, BMI, and smoking—strongly supports the foundational claim that androgen signaling actively drives pain resolution [10]. The statistical relationship between testosterone and pain resolution appears consistent across both sexes, suggesting that the androgen-driven protective effect is not restricted to males, though absolute hormone levels remain the primary driver of the observed sex dimorphism in population outcomes [11].

We acknowledge that the UKBB phenotype represents a ‘static’ snapshot of a dynamic process. However, by contrasting individuals with recently resolved pain against those with persistent chronic pain, we capture the population-level result of the resolution trajectory. While this lacks the temporal precision of the AURORA trauma cohort [12], its scale provides the statistical power to identify independent hormonal contributors like testosterone that are otherwise masked in smaller, more homogeneous studies.

### Clinical Implications and Future Directions

These findings present distinct strategic avenues for precision medicine and companion diagnostics in chronic pain management. Because total monocyte count is heavily confounded by generalized inflammation [13], future research should pivot toward identifying genetic variants that alter monocyte activation states. For example, investigating whether single nucleotide polymorphisms (SNPs) in localized innate immune receptors—such as Toll-like receptors (TLR7/8)—influence the propensity of monocytes to adopt a resolution-promoting phenotype could yield highly precise companion diagnostics. Such stratification tools could identify patients at severe risk of failing to resolve acute traumatic pain, facilitating early, targeted therapeutic intervention.

Because total peripheral monocyte counts are heavily confounded by metabolic factors (BMI, smoking) and systemic IL-10, the path toward a viable companion diagnostic lies in identifying the **genetic potential** for resolution. Genotyping for variants that modulate monocyte activation (e.g., TLR7/8 polymorphisms) may identify individuals who possess the cellular machinery to resolve pain, even if that machinery is not currently ‘active’ in their peripheral blood at the time of testing.

The divergence between our IL-10 findings and those of the AURORA study likely reflects the ‘acute-stress’ versus ‘baseline-state’ measurement. AURORA captured IL-10 during the immediate post-traumatic inflammatory surge [12], whereas UKBB captures a baseline physiological state. Our results suggest that IL-10 is a marker of **active** resolution rather than a baseline predictor of **future** resolution.

## Limitations

Several limitations of the present study warrant consideration, particularly when interpreting our findings alongside the mechanistic framework established by Sim et al. (2026).

### 1. Systemic vs. Localized Neuroimmune Signaling

The most significant conceptual limitation is the difference in biological compartments. Sim et al. demonstrated that IL-10^+^ monocytes drive pain resolution through localized signaling within inflamed tissue (e.g., the skin) directly to IL-10R1^+^ sensory neurons. In contrast, our study utilized total peripheral blood monocyte counts and systemic plasma IL-10 levels. It is highly probable that the specialized, resolution-promoting immune activity is compartmentalized at the site of injury and is too localized or transient to be accurately captured by systemic biomarkers in a broad population survey. Therefore, our null findings for IL-10 and monocytes do not necessarily refute the local mechanism but rather highlight the difficulty of measuring these events in the peripheral circulation.

### 2. Temporal Phenotyping and Measurement Timing

While the AURORA study measured biomarkers during the acute post-traumatic phase, the UK Biobank provides a baseline “snapshot.” Although we utilized a longitudinal transition model to mirror the Aurora 84-day resolution window, the mean interval between UKB assessments was significantly longer (1,579 days). This temporal gap may miss the critical resolution window where transient IL-10 spikes or specialized monocyte subsets are most influential. Our findings suggest that while these biomarkers may be potent predictors of *acute* resolution, they may not persist as stable markers of long-term recovery in a general middle-aged population.

### 3. Etiological Heterogeneity

The UK Biobank pain phenotype is highly heterogeneous, encompassing diverse etiologies including mechanical, neuropathic, and multi-site inflammatory pain. The biological drivers of resolution may vary significantly across these different pain types, potentially diluting the effect size of the androgen-monocyte axis which was primarily characterized in inflammatory and traumatic contexts.

The UK Biobank pain phenotype is unlike controlled inflammatory models and CFA injection. Complete Freund’s Adjuvant (CFA) injection in mice, often used for antibody production or inflammation modeling, involves injecting an emulsion of antigen, paraffin oil, and Mycobacterium tuberculosis. UK Biobank is also unlike the traumatic injury cohorts of AURORA.

### 4. Confounding by Metabolic Health

Our longitudinal results emphasize that in real-world human populations, pro-inflammatory lifestyle factors such as high BMI and smoking are dominant predictors of pain persistence. These factors likely “mask” the native neuroimmune resolution machinery. Future studies should prioritize the identification of genetic variants (e.g., SNPs in TLR7/8 [14, 15]) that govern the *potential* for monocyte activation, as these stable genetic traits may serve as more reliable predictors than highly variable systemic cell counts or protein levels.

### 5. Discrepancy between clinical observation windows and biological resolution phases

A primary critique of epidemiological pain research is the discrepancy between clinical observation windows and biological resolution phases. While preclinical models often define resolution within days, our study utilizes the UK Biobank’s multi-year follow-up to identify a phenotype of **biological persistence**.

The fact that LPO and RTN4R remain significant predictors over an average interval of 4.3 years suggests that these are not merely transient markers of acute inflammation, but rather signatures of a ‘locked’ physiological state. We propose that LPO-driven oxidative damage may lead to permanent or semi-permanent changes in the localized tissue environment, effectively ‘priming’ the nervous system for chronic pain. The independence of these markers from systemic monocyte counts in both sexes further supports the hypothesis that the functional activity of these cells—rather than their absolute quantity—is the determinant of whether an individual enters a state of long-term pain persistence.

### 6. 4.3-year interval between baseline and follow-up

We acknowledge that the 4.3-year interval between baseline and follow-up in the UK Biobank does not capture the immediate ‘resolution window’ (days-to-weeks) typically studied in preclinical models. Therefore, our results should be interpreted as identifying markers of **long-term pain persistence** rather than the acute phase of biological resolution.

## Conclusion

Our study demonstrates that the transition from acute to chronic pain is not merely a consequence of systemic immune or hormonal status, but is driven by specific, high-precision molecular signatures. By screening nearly 3,000 proteins, we identified **Lactoperoxidase (LPO)** and **Nogo Receptor (RTN4R)** as dominant independent predictors of pain chronification.

The finding that LPO—a marker of activated myeloid oxidative activity—supersedes the predictive power of absolute monocyte counts suggests that the *functional state* of immune cells is more critical to pain resolution than their systemic quantity. Furthermore, the association of RTN4R points to a structural failure in neural repair as a key barrier to recovery.

While our population-level data continues to support the protective role of the androgen-pain axis, these proteomic results suggest that testosterone likely operates by modulating these specific oxidative and neural-inhibitory pathways. Ultimately, these results shift the focus of pain research from broad systemic inflammation to localized molecular precision, identifying LPO and RTN4R as novel candidates for further mechanistic investigation for preventing the development of chronic pain.

## Data Availability

The data analyzed in this study were obtained from the UK Biobank (UKBB) Resource under Application Number 57245. The raw datasets, including the UKB Pharma Proteomics Project (UKB-PPP) Olink data, hematology, and touchscreen questionnaires, are available to bona fide researchers upon application to the UK Biobank (www.ukbiobank.ac.uk/enable-your-research/apply-for-access).

## References

[1] Zhang YL, Wu XC, Chen XY, Gao F, Wang J (2025) Global and regional burden, temporal trends, and projections of chronic pain from 1990 to 2032, and its association with cardiovascular diseases: analyses based on global burden of diseases study 2021. Front Public Health 13, 1636949.

2. [2] Barreto MCA, Ávila MA, Cartes-Velásquez R, de Castro SS (2025) The Burden of Chronic Pain on Women: A Secondary Analysis of Data From the National Study on Disability (ENDISC) in Chile. Eur J Pain 29, e70080.

[3] Fang XX, Zhai MN, Zhu M, He C, Wang H, Wang J, Zhang ZJ (2023) Inflammation in pathogenesis of chronic pain: Foe and friend. Mol Pain 19, 17448069231178176.

[4] Hakim S, Jain A, Woolf CJ (2024) Immune drivers of pain resolution and protection. Nature Immunology 25, 2200–2208.

[5] Chen Y, Wu X, Li J, Ren Y, Miao H, Zhai X, Huang C, Chen X (2025) The mechanisms of specialized pro-resolving mediators in pain relief: neuro-immune and neuroglial regulations. Frontiers in Immunology Volume 16 **-** 2025.

6. [6] Sim J, O’Guin E, Sugimoto C, Laumet S, Monahan K, Bernard MP, McLean SA, Albertorio-Sáez LM, Zhao Y, Ramakrishnan H, de Souza S, Eller OC, Smoyer CJ, Baumbauer KM, Mack M, Folger JK, Robison AJ, Linnstaedt SD, Laumet G (2026) Monocyte-derived IL-10 drives sex differences in pain duration. Science Immunology 11, eadx0292.

[7] Sudlow C, Gallacher J, Allen N, Beral V, Burton P, Danesh J, Downey P, Elliott P, Green J, Landray M, Liu B, Matthews P, Ong G, Pell J, Silman A, Young A, Sprosen T, Peakman T, Collins R (2015) UK biobank: an open access resource for identifying the causes of a wide range of complex diseases of middle and old age. PLoS Med 12, e1001779.

[8] Eldjarn GH, Ferkingstad E, Lund SH, Helgason H, Magnusson OT, Gunnarsdottir K, Olafsdottir TA, Halldorsson BV, Olason PI, Zink F, Gudjonsson SA, Sveinbjornsson G, Magnusson MI, Helgason A, Oddsson A, Halldorsson GH, Magnusson MK, Saevarsdottir S, Eiriksdottir T, Masson G, Stefansson H, Jonsdottir I, Holm H, Rafnar T, Melsted P, Saemundsdottir J, Norddahl GL, Thorleifsson G, Ulfarsson MO, Gudbjartsson DF, Thorsteinsdottir U, Sulem P, Stefansson K (2023) Large-scale plasma proteomics comparisons through genetics and disease associations. Nature 622, 348–358.

[9] Martinez-Moreno JM, Llamas-Urbano A, Barbarroja N, Perez-Sanchez C (2025) Proteomics by qPCR Using the Proximity Extension Assay (PEA). Methods Mol Biol 2929, 129–142.

[10] Saika F, Uta D, Fukazawa Y, Hino Y, Hatano Y, Kishioka S, Nawa H, Hino S, Suzuki K, Kiguchi N (2025) Androgen receptors expressed in the primary sensory neurons regulate mechanical pain sensitivity. Pain 166, e746–e757.

[11] Alimoradian A, Abbaszadeh F, Jorjani M (2024) Testosterone signaling pathways for reducing neuropathic pain in a rat model of spinothalamic tract lesion. Iran J Basic Med Sci 27, 1417–1422.

[12] McLean SA, Ressler K, Koenen KC, Neylan T, Germine L, Jovanovic T, Clifford GD, Zeng D, An X, Linnstaedt S, Beaudoin F, House S, Bollen KA, Musey P, Hendry P, Jones CW, Lewandowski C, Swor R, Datner E, Mohiuddin K, Stevens JS, Storrow A, Kurz MC, McGrath ME, Fermann GJ, Hudak LA, Gentile N, Chang AM, Peak DA, Pascual JL, Seamon MJ, Sergot P, Peacock WF, Diercks D, Sanchez LD, Rathlev N, Domeier R, Haran JP, Pearson C, Murty VP, Insel TR, Dagum P, Onnela JP, Bruce SE, Gaynes BN, Joormann J, Miller MW, Pietrzak RH, Buysse DJ, Pizzagalli DA, Rauch SL, Harte SE, Young LJ, Barch DM, Lebois LAM, van Rooij SJH, Luna B, Smoller JW, Dougherty RF, Pace TWW, Binder E, Sheridan JF, Elliott JM, Basu A, Fromer M, Parlikar T, Zaslavsky AM, Kessler R (2020) The AURORA Study: a longitudinal, multimodal library of brain biology and function after traumatic stress exposure. Mol Psychiatry 25, 283–296.

[13] Kim JH, Lee YJ, Park B (2019) Higher monocyte count with normal white blood cell count is positively associated with 10-year cardiovascular disease risk determined by Framingham risk score among community-dwelling Korean individuals. Medicine (Baltimore) 98, e15340.

[14] Gotay WJP, Maciel MSC, Rodrigues RO, Cardoso CC, Oliveira CN, Montenegro AFL, Yaochite JNU (2025) X-linked polymorphisms in TLR7 and TLR8 genes are associated with protection against Chikungunya fever. Mem Inst Oswaldo Cruz 120, e230224.

[15] Hamerman JA, Barton GM (2024) The path ahead for understanding Toll-like receptor-driven systemic autoimmunity. Curr Opin Immunol 91, 102482.

